# Disentangling the Shared and Differential Genetic Architecture Between COVID-19 and Other Respiratory Disorders: A Multi-Omics Genome-Wide Analysis

**DOI:** 10.64898/2026.03.21.26348591

**Authors:** Xiao Xue, Yu-Ping Lin, Yaning Feng, Hon-Cheong So

## Abstract

**Background:** A bidirectional relationship has been observed between COVID-19 and respiratory disorders, where respiratory comorbidities increase severity and COVID-19 induces respiratory sequelae. The underlying biological and genetic mechanisms remain unclear. While previous studies have identified overlapping genetic loci, few have systematically disentangled the genetic factors shared between these conditions versus those specific to COVID-19, particularly at a multi-omics level.

**Methods:** We developed and applied a unified analytical framework to compare three COVID-19 phenotypes with eight respiratory disorders (including asthma, COPD, IPF, and pneumonia). Utilizing the cofdr method for shared genetic signal analysis and DDx/mtCOJO for differentiation, we integrated genome-wide association statistics with multi-omics data (transcriptome, splicing, and proteome). This approach allowed for the simultaneous identification of shared genetic signals (concordant or discordant) and disease-specific variants across expression (TWAS), alternative splicing (spTWAS), and protein abundance (PWAS).

**Results:** We delineated a comprehensive atlas of 214 differential and numerous shared loci across 24 pairwise comparisons. The shared genetic architecture was characterized by pleiotropic effects in genes such as *ATP11A* (exhibiting opposing effects in COVID-19 vs. IPF) and *GSDMB* (shared with COPD). Crucially, differentiation analysis revealed that severe COVID-19 is genetically distinct from other respiratory infections (e.g., pneumonia and influenza) through dysregulated Type I/III interferon signaling and specific defects in alveolar epithelial and macrophage function, as well as GM-CSF/surfactant metabolism pathways. These findings provide direct genetic evidence supporting the use of GM-CSF modulators and interferon-lambda for COVID-19 treatment, therapies that have already entered clinical trials. Furthermore, multi-trait conditional analysis prioritized *FYCO1* and *HCN3* as potential COVID-19-specific risk genes. Splicing analysis underscored the critical role of alternative splicing in both shared and differential architectures, highlighting *IFNAR2* isoform regulation as a key discriminator between COVID-19 and other respiratory traits.

**Conclusion:** This study provides the first genome-wide, multi-omics map revealing the shared and differential genetic landscapes of COVID-19 and other respiratory phenotypes. By uncovering specific molecular mechanisms that distinguish COVID-19 pathology, specifically involving surfactant homeostasis and interferon pathways, our findings offer novel insights for targeted drug repurposing and precision risk stratification.

## Introduction

Coronavirus disease 2019 (COVID-19), caused by severe acute respiratory syndrome coronavirus type 2 (SARS-CoV-2), has greatly impacted global health, affecting hundreds of millions of individuals and causing millions of deaths worldwide. An important feature of COVID-19 is the remarkable heterogeneity in clinical outcomes. While many experience mild symptoms, others progress to severe disease. Understanding the factors that determine this heterogeneity is of paramount importance for risk stratification, early intervention, and the development of targeted therapeutics.

Epidemiological studies have consistently identified pre-existing *respiratory disorders* as major risk factors for adverse COVID-19 outcomes. Chronic obstructive pulmonary disease (COPD) is associated with a substantially increased risk of severe COVID-19, hospitalization, and mortality^1, 2^. Similarly, patients with interstitial lung disease (ILD) and idiopathic pulmonary fibrosis (IPF) experience poor outcomes following SARS-CoV-2 infection^3, 4^. Prior influenza infection (especially within one year) has also been linked to increased COVID-19 severity^5^. The relationship between asthma and COVID-19 severity is more complex, with some studies reporting increased risk of hospitalization^6^ while others suggest protective effects^7, 8^, a discrepancy that remains incompletely understood.

Beyond serving as risk factors for acute infection, respiratory comorbidities may also emerge as sequelae of COVID-19, with growing evidence that COVID-19 survivors experience long-lasting pulmonary impairment^9, 10^, lung fibrosis^9^, and increased susceptibility to subsequent respiratory disorders including COPD or COPD exacerbation^11^.

### Importance of understanding shared and differential genetic architecture

The bidirectional relationships between COVID-19 and other respiratory disorders raise fundamental questions about the underlying biological mechanisms. Shared genetic factors and pathophysiology with other respiratory disorders may explain why certain pulmonary conditions serve as risk factors for severe COVID-19 and/or develop as sequelae. From a clinical perspective, identifying shared genetic factors could inform risk prediction for severe infection or sequelae, enable targeted screening of high-risk individuals, and reveal common therapeutic targets for comorbid diseases.

Conversely, the observation that only a subset of patients with respiratory comorbidities develops severe COVID-19, and that only some survivors experience pulmonary sequelae, suggests the existence of disease-specific genetic factors. In addition, studies have suggested that COVID-19 is associated with higher mortalities compared to influenza^12–14^, suggesting that unique pathological mechanisms may underlie COVID-19. Identifying COVID-19-specific variants could illuminate unique aspects of SARS-CoV-2 pathogenesis and reveal therapeutic targets that could be modulated without affecting other respiratory conditions. Understanding both the shared and differential genetic architecture between COVID-19 and other respiratory disorders is therefore of substantial scientific and clinical importance.

### Previous relevant studies and limitations

Genome-wide association studies (GWAS) have made substantial progress in identifying genetic variants associated with COVID-19 susceptibility and severity. The COVID-19 Host Genetics Initiative (HGI) has identified numerous risk loci through international collaboration^15^. Similarly, well-powered GWAS have been conducted for respiratory disorders including asthma, COPD, IPF, and pneumonia, identifying hundreds of associated loci^16–20^. Several studies have begun to explore genetic overlap between COVID-19 and respiratory conditions^21–24^. For example, a cross-trait meta-analysis identified shared loci between COVID-19 hospitalization and asthma, including variants within *ABO* and *ATXN2*^22^. Another colocalization analysis detected a shared signal at rs12610495 between severe COVID-19 and IPF, implicating an expression quantitative trait locus (eQTL) for DPP9 in lung tissue^21, 25^. Genetic correlation analyses have documented significant overlap between COVID-19 and various respiratory phenotypes^25, 26^.

Despite the progress, several important limitations are present for existing cross-trait genetic studies of COVID-19 and respiratory disorders. **First,** most studies have focused on identifying shared genetic effects through colocalization or genetic correlation, *without systematically examining disease-specific (differentially associated) variants or those with opposing effects between conditions*. This one-sided approach provides an incomplete picture of cross-trait genetic architecture. **Second**, existing studies have largely operated at the SNP level, with limited *integration of multi-omics data* to reveal functional mechanisms. While some studies have incorporated expression QTL data^27^, few have simultaneously examined splicing QTL (sQTL) and protein QTL (pQTL), despite growing evidence that alternative splicing and post-transcriptional regulation play important roles in respiratory disease pathogenesis^28–32^. **Third**, most previous analyses have examined only pairwise comparisons between COVID-19 and a single respiratory trait, rather than simultaneously analyzing multiple conditions to identify variants with consistent versus heterogeneous effects *across the respiratory disease spectrum*. **Fourth**, very few studies employ a *genome-wide scan* approach to examine shared or differential SNPs/genes between disorders across the entire genome. Some studies focused on a restricted set of genetic variants, for example those passing genome-wide significance in COVID-19 and tested whether they are implicated in other disorders^33^. By ignoring the broader polygenic architecture, these studies likely miss smaller-effect variants that contribute I to shared or divergent genetic risk, thereby limiting the discovery of novel biological pathways.

### Study goals and analytical approach

To address these gaps, we presented and applied a comprehensive framework to systematically uncover both shared and specific genetic factors underlying COVID-19 and other respiratory disorders through integration of large-scale genetic data with multi-omics resources. Our framework has several key features that distinguish it from previous approaches.

First, we integrate shared genetic signal/colocalization analysis with differentiation analysis within a unified pipeline. Methods such as cofdr, gwas-pw and hyprcoloc identify variants with *shared* effects across traits, while *differentiation* methods (DDx, CC-GWAS, mtCOJO) identify variants with heterogeneous or disease-specific effects. Crucially, rather than relying on qualitative assessments, all selected methods provide statistical significance measures through *p*-values or posterior probabilities. By applying both approaches, we can comprehensively characterize genetic variants as shared with concordant effects, shared with discordant effects, specific to one condition, or not associated with either condition.

Second, we extend cross-trait analysis beyond SNP-level associations to gene-level effects derived from multiple ‘omics’ layers. By considering gene-level statistics from MAGMA (positional mapping), TWAS (expression), spTWAS (splicing), and PWAS (protein abundance), we identified potential shared and differential genes and then characterize whether they influence the disorders through effects on gene expression, alternative splicing, and/or protein levels.

Third, we also attempt to prioritize and validate our findings through convergent evidence from multiple independent methods, thereby reducing false discoveries that may arise from method-specific assumptions or biases.

We applied this framework to compare GWAS data on three COVID-19 phenotypes (severe/critical COVID-19 [A2], hospitalized COVID-19 [B2], and reported SARS-CoV-2 infection [C2]) with 8 respiratory Hi disorders (asthma, ILD, IPF, COPD, pneumonia, bacterial pneumonia, viral pneumonia, and influenza), encompassing 24 pairwise comparisons and 3 multi-trait comparisons. Shared and differentially associated variants and genes were identified. Enriched gene sets, tissues, and cell types were also detected.

Importantly, our analytic framework has been made openly available on GitHub, and we believe this will serve as a useful resource for researchers in human genetics.

To our knowledge, this is the first study to employ a *genome-wide* and *multi-omics* framework to map the shared and divergent genetic landscapes between COVID-19 and a comprehensive spectrum of respiratory disorders. Overall, our results provide a comprehensive atlas of shared and differential genetic architecture between COVID-19 and other respiratory diseases, with implications for understanding disease mechanisms and informing personalized approaches to prevention and treatment.

## Methods

### GWAS summary statistics

#### GWAS data harmonization

We leveraged data from the COVID-19 Host Genetics Initiative (COVID-19 HGI Release7)^15^ comparing genetic structures between COVID-19 with other related respiratory conditions. GWAS summary statistics were collected for 8 respiratory disorders related to COVID-19^34^. GWAS datasets were formatted and harmonized using MungeSumstats^35^, with default parameters. Note that *MungeSumstats* ensures consistency of allele assignment and direction of effects across studies. Summary statistics were aligned to GRCh37, and only bi-allelic SNPs with MAF≥0.01 present in the reference file (dbSNP144) were preserved.

#### GWAS meta-analysis

A meta-analysis of pneumonia summary statistics was performed using METAL^36^ between PanUKBB (phecode 480) and Finngen (J10 pneumonia) samples. To impute unmeasured SNPs, meta-analyzed variants present in both cohorts were utilized as input for DIST^37^ using the 1000 Genome EUR reference panel. Follow-up studies were conducted on the imputed common variants (INFO>0.3). Using the above workflow, we also generated meta-analyzed GWAS for bacterial pneumonia (PanUKBB phecode 480.1; Finngen J10 bacterial pneumonia), viral pneumonia (PanUKBB phecode 480.2; Finngen J10 viral pneumonia), and influenza (PanUKBB phecode 481; Finngen influenza).

### SNP-based decorrelation

A decorrelated Z-score matrix was generated to minimize the effect of sample overlap, based on the method described in ref ^39^:

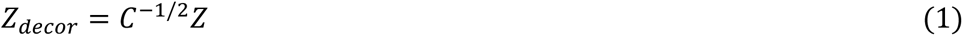

where C is a matrix with ones at the diagonal and the LDSC intercept^40^ as off-diagonal elements representing sample overlap, and the matrix **Z** contains the original Z-scores. For GWAS with unreliable LDSC estimates due to small sample sizes, sample overlap was corrected using the correlation between Z-scores of putatively null SNPs, following the approach used by the software METAL for overlapping samples^36, 41^. First, strongly associated loci were removed applying a cutoff value for absolute Z-scores (|Z|<2). Second, the covariance matrix was derived by fitting “mle.tmvnorm“^42^ with truncated Z-scores, and the corresponding correlation between the two traits was calculated. As shown in Supplementary Table 1, most of the traits studied showed only minimal or small sample overlap.

### Shared genetic architecture at the SNP level

#### Pairwise shared-variant analysis using cofdr

To identify genetic variants that jointly contribute to the risk of COVID-19 and other related traits, the ‘cofdr’ algorithm (proposed in ref^43^) was applied with a decorrelated Z-score matrix as input. Briefly, a four-group mixture model was constructed to determine whether a genetic variant is associated with trait1, trait2, both traits, or neither trait1 nor trait2.

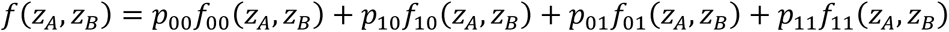

The likelihood function was maximized with the EM algorithm to estimate the posterior probability that an SNP is shared by two traits (defined as posterior probability≥0.9 in this study), associated with trait 1 (or trait 2) only, and associated with neither trait 1 nor trait 2. These estimates can be considered as a two-dimensional extension of the standard local false discovery rate^44^. Here we allow a highly flexible distribution of the non-null effects, based on a continuous mixture model. The distribution of *z* under the alternative is assumed to follow

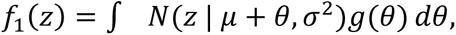

where *g*(*θ*) is a flexible distribution for the non-null effects. We employed the “prfdr” algorithm proposed by Scott et al.^45^ to estimate the alternative density. The bivariate density is estimated by the product of the marginal densities (after the decorrelation step). The method can be readily extended to more than two traits; for example, an 8-component mixture model can be constructed for three traits. For details, please refer to ref^43^ and the supplementary information therein.

Cofdr also conducted likelihood ratio test for the existence of pleiotropy at a genome-wide level, testing the null hypothesis *H*_0_ : *π*_11_ = *π*_1⋅_× *π*_⋅1_ (i.e., independence of association status across traits), where π₁₁ denotes the proportion of SNPs associated with both traits, and π₁· and π·₁ denote the marginal proportions associated with each trait, respectively.

While cofdr is designed to identify shared or pleiotropic genetic signals, methods such as gwas-pw, coloc, and hyprcoloc represent alternative approaches that detect shared causal variants based on linkage disequilibrium (LD) structure. We employ a hybrid strategy here, as these two classes of methods are complementary. Cofdr is highly scalable to millions of SNPs and multiple traits, enabling efficient genome-wide scanning for shared variants. In contrast, coloc-type algorithms typically focus on individual loci of interest.

Notably, cofdr does not require an LD reference panel, which reduces issues arising from ancestry mismatch between the reference panel and the study population. It also provides a likelihood ratio test for genetic overlap at a genome-wide level, and requires only p-values (or z-scores), the most commonly available summary statistics. Consequently, cofdr can handle gene-level statistics (such as those from MAGMA, TWAS, SpTWAS, or PWAS) as input and estimate the posterior probability of shared gene-level signals. As coloc-type algorithms are primarily designed for SNP-level data, there is currently a lack of studies investigating shared signals at the gene level across multiple molecular layers.

On the other hand, coloc-type algorithms are specifically suited to determine whether two traits share the same *causal* variant at a specific locus while explicitly accounting for LD information and therefore serve a complementary objective.

#### Prioritization of shared genetic variants using gwas-pw and hyprcoloc

Shared SNPs identified from cofdr were further prioritized with gwas-pw^46^ and hyprcoloc^47^. Gwas-pw computes the posterior probability of association (PPA) for pairwise comparisons that LD-independent genomic regions^48^ are implicated in only one trait (models 1 and 2), both traits (model 3), or both traits with different variations (model 4). Correlations estimated by “mle.tmvnorm” or LDSC intercepts were used as correction factors in overlapping cohorts. A putative causal variant can then be identified by estimating the PPA for each SNP within a predefined LD-independent segment. Genomic regions with PPA ≥ 0.8 for model 3 were considered pleiotropic; within such regions, the SNP with the highest PPA (≥ 0.5) was designated as the shared causal variant. The same predefined LD-independent blocks within European populations were used as testing regions for hyprcoloc, a Bayesian divisive clustering algorithm. Hyprcoloc estimates the posterior probability of traits colocalizing within a given region, identifies the putative causal variant in this region, and the proportion of the posterior probability explained by that variant.

#### Multi-trait shared loci analysis

Moreover, a multi-trait analysis was conducted to identify shared signals using cofdr. Specifically, we explored shared genetic architecture between COVID-19 and multiple respiratory disorders simultaneously, focusing on traits that shared at least one validated locus with COVID-19 in pairwise comparisons. Hyprcoloc was applied for further prioritization and validation of the results.

### Shared signals at the gene level - a multi-omics approach

#### Gene-based shared signals based on MAGMA (MAGMA-cofdr)

To identify genes shared between COVID-19 and other related medical conditions, we performed gene-based analysis using MAGMA^49^. We first decorrelated SNP-based P values of involved traits to minimize the influence of sample overlap. We captured variants within the gene body only. Computed gene-based P values were converted into two-sided Z-values and input to cofdr. A posterior probability threshold of ≥ 0.9 was used to identify “shared” genes.

#### Gene-based shared signals from TWAS/spTWAS (TWAS/spTWAS-cofdr)

Second, we investigated shared genetically regulated effects on gene expression and alternative splicing across traits using transcriptome-wide association studies (TWAS) and splicing TWAS (spTWAS). We conducted TWAS/spTWAS using decorrelated GWAS data to obtain gene-based Z-scores as input for cofdr. We employed MASHR-based eQTL/sQTL models^50^ pre-trained on GTEx v8 data (49 tissues, N=838)^51^. Prediction models for each tissue were integrated with decorrelated GWAS statistics using S-PrediXcan^52^, and association statistics across all tissues were combined using S-MultiXcan^53^.

Similar to above, we applied a posterior probability (π_TT) threshold of 0.9. For splicing analysis, colocalized intron IDs discovered by spTWAS-cofdr were mapped to gene IDs using annotations from leafcutter^54^. Meanwhile, for a gene containing multiple colocalized intron junctions and thus multiple π_TT values, we only report the largest one for that gene.

#### Gene-based shared signals from PWAS (PWAS-cofdr)

Similarly, we conducted a protein-wide association study (PWAS) to determine proteins that are jointly related to COVID-19 and other related respiratory disorders. In total, 1348 predictive models of plasma proteins fitted from 7213 European Americans (EA)^55^ were used for downstream PWAS using the tool FUSION^56^, based on harmonized decorrelated GWAS summary statistics. Significant shared proteins were defined as those with a posterior probability from cofdr greater than 0.8 (a slightly more liberal threshold was employed here given the smaller number of testable proteins in the PWAS-cоfdr analysis).

### Differentiation analysis at the SNP level

#### Pairwise differentiation analysis using DDx

To identify genomic loci with divergent effects between COVID-19 and other respiratory traits, we performed case–case GWAS using the “DDx” methodology we recently proposed (please see ref^57^ for details of the method). In essence, this approach treats one trait as the case and the other as the control and performs a GWAS. Sample overlap is also accounted for. SNPs with P<5e-8 in the derived case-case GWAS were considered significant for differential effects.

#### Validation of differentially associated variants using CC-GWAS

CC-GWAS^58^ was applied to validate the significant variants identified by DDx. Specifically, the algorithm computes case-case associations of two different disorders based on their respective case-control GWAS results. CC-GWAS combines two components, CC-GWAS_OLS_ and CC-GWAS_Exact_, to control type 1 error, and SNPs with OLS_pval<5e-8 and Exact_pval<1e-4 were considered statistically significant.

### Multi-trait conditional analysis using mtCOJO

Whereas DDx performs a case-case comparison between any two traits, multi-trait conditional and joint analysis (mtCOJO)^59^ enables GWAS of a target phenotype Y conditioned on one or more genetically correlated phenotypes X. If X is genetically correlated with Y, SNP associations may become more or less significant after conditioning. mtCOJO can sometimes reveal genome-wide significant SNPs that are not significant in the unadjusted GWAS. Furthermore, mtCOJO allows simultaneous conditioning on multiple traits.

We performed multivariate conditional analyses using mtCOJO to condition the effect of each SNP on COVID-19 upon genetically correlated respiratory traits. Genome-wide significant SNPs were identified in the conditional analysis at P<5e-8. MAGMA gene and gene-set analyses were conducted to identify genes and gene sets associated with the conditioned COVID-19 (*MAGMA-mtCOJO)*. Tissue and cell-type specificity analyses were also performed using FUMA. Moreover, we applied TWAS, spTWAS, and PWAS to the conditioned GWAS to test the effects of disorder-specific variants on gene expression, splicing, and protein levels (TWAS-mtCOJO, spTWAS-mtCOJO, and PWAS-mtCOJO, respectively). Significant genes were identified at FDR < 0.01.

### Differentiation genetic architecture at the gene level

Gene-level differentiation analysis was first performed by applying MAGMA to DDx-derived case-case GWAS (MAGMA-DDx), which involves physically mapping SNPs to genes. Next, to identify genes with divergent expression, splicing, or protein levels between traits, DDx-derived GWAS statistics were integrated with eQTL, sQTL, and pQTL data (TWAS-DDx, spTWAS-DDx, and PWAS-DDx, respectively). All differential genes were further compared with findings based on CC-GWAS_OLS_ case-case effect sizes. Due to the relatively large number of significant results, we mainly present findings with FDR<0.01.

### Post-GWAS functional annotation

Functional characterization and prioritization of the identified genomic risk loci were performed by FUMA^60^. We utilized the SNP2GENE module for functional annotation and GENE2FUNC for pathway enrichment analysis of colocalized genes. To identify tissues and cell types associated with differential effects, we conducted MAGMA gene-property analyses by integrating case-case GWAS with GTEx v8 tissue expression data and 12 single-cell RNA-seq datasets.

### Phenome-wide Association (PheWAS) analysis

The phenotypic associations of significant genes were investigated in the GWAS Atlas^61^ to characterize their pleiotropic effects. The GWAS Atlas provides an integrated database of 4,756 GWAS covering 3,302 unique traits across 28 domains. Significant phenotypes classified into respiratory, cardiovascular, and immunological domains were extracted using Bonferroni-corrected p-values < 0.05/number of tested GWASs.

## Results

### Shared genetic signals

#### Pairwise shared signals at the SNP-level

SNP-level analyses were conducted to find shared genomic risk loci between pairs of traits. In A2-related (severe/critical COVID-19) comparisons, cоfdr identified 113 risk loci, of which 14 were prioritized/validated by both gwas-pw and hyprcoloc. In B2-related (hospitalized COVID-19) analyses, 119 loci were identified with 11 validated, and in C2-related (reported COVID-19) analyses, 29 loci were identified with 5 validated (Table 2). Among all validated loci, 17 had p<5×10⁻⁶ in the original GWAS for both traits, with 9 showing concordant allelic effects and 8 showing opposing effects (Table 3).

**Table 1.**
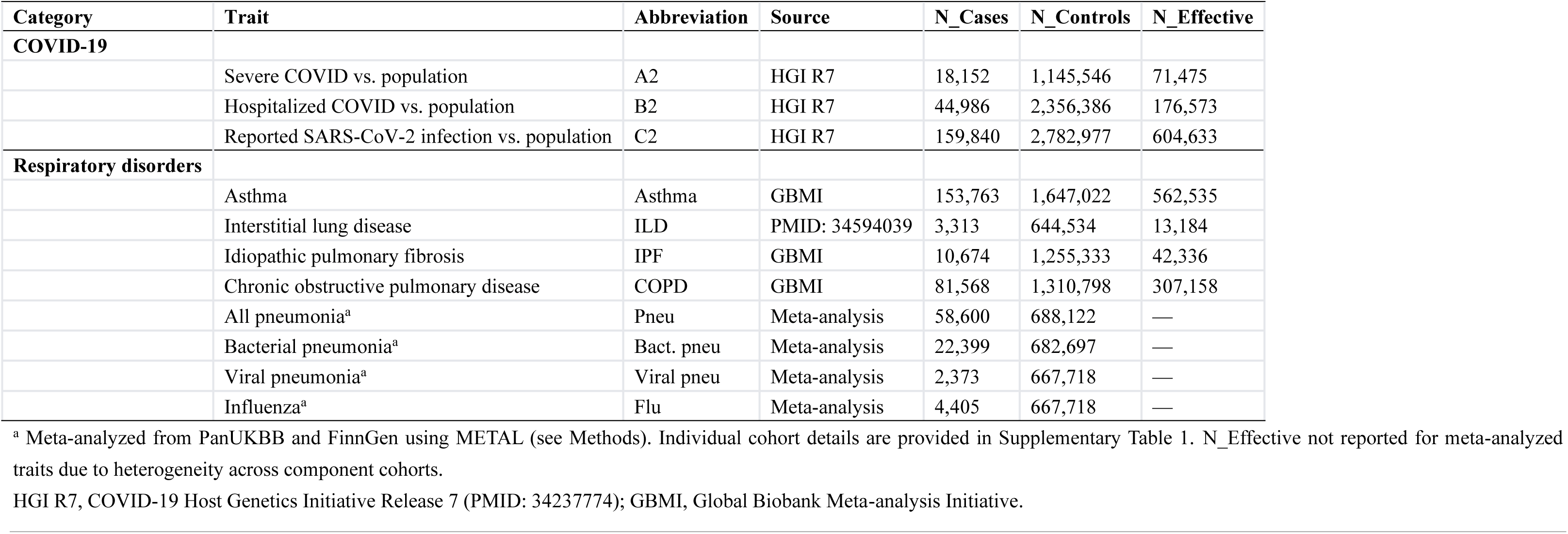
Summary of analyzed GWAS.

| Category | Trait | Abbreviation | Source | N_Cases | N_Controls | N_Effective |
| --- | --- | --- | --- | --- | --- | --- |
| COVID-19 |  |  |  |  |  |  |
|  | Severe COVID vs. population | A2 | HGI R7 | 18,152 | 1,145,546 | 71,475 |
|  | Hospitalized COVID vs. population | B2 | HGI R7 | 44,986 | 2,356,386 | 176,573 |
|  | Reported SARS-CoV-2 infection vs. population | C2 | HGI R7 | 159,840 | 2,782,977 | 604,633 |
| Respiratory disorders |  |  |  |  |  |  |
|  | Asthma | Asthma | GBMI | 153,763 | 1,647,022 | 562,535 |
|  | Interstitial lung disease | ILD | PMID: 34594039 | 3,313 | 644,534 | 13,184 |
|  | Idiopathic pulmonary fibrosis | IPF | GBMI | 10,674 | 1,255,333 | 42,336 |
|  | Chronic obstructive pulmonary disease | COPD | GBMI | 81,568 | 1,310,798 | 307,158 |
|  | All pneumonia<sup>a</sup> | Pneu | Meta-analysis | 58,600 | 688,122 | — |
|  | Bacterial pneumonia<sup>a</sup> | Bact. pneu | Meta-analysis | 22,399 | 682,697 | — |
|  | Viral pneumonia<sup>a</sup> | Viral pneu | Meta-analysis | 2,373 | 667,718 | — |
|  | Influenza<sup>a</sup> | Flu | Meta-analysis | 4,405 | 667,718 | — |

<sup>a</sup> Meta-analyzed from PanUKBB and FinnGen using METAL (see Methods). Individual cohort details are provided in Supplementary Table 1. N\_Effective not reported for meta-analyzed traits due to heterogeneity across component cohorts.
HGI R7, COVID-19 Host Genetics Initiative Release 7 (PMID: 34237774); GBMI, Global Biobank Meta-analysis Initiative.

**Table 2.**
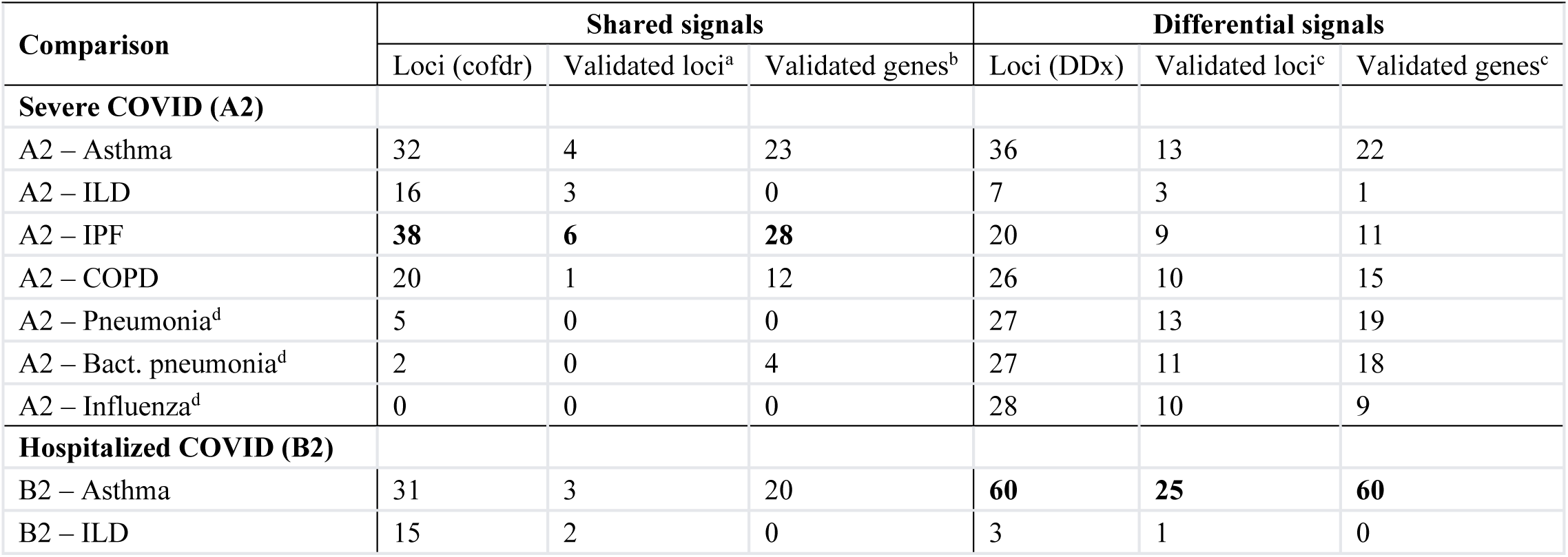
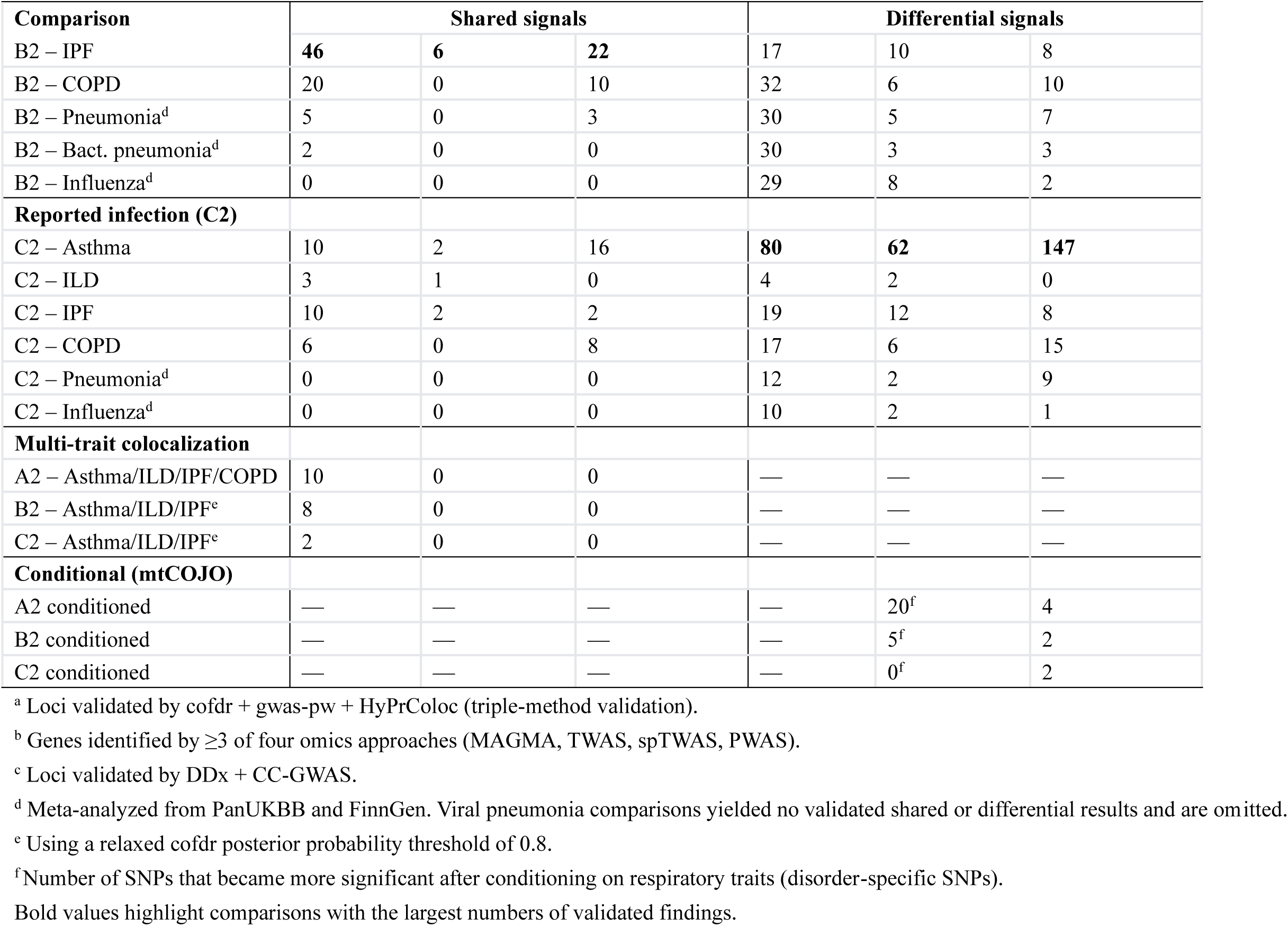
Overview of shared and differential findings across all analyses.

**Table 3.** Validated shared loci between COVID-19 and respiratory disorders.

| Comparison | Lead SNP | Chr:Pos | EA | $\beta_{\text{COVID}}$ | $\beta_{\text{Resp}}$ | P_COVID | P_Resp | Dir.<sup>a</sup> | Nearest gene(s) | PheWAS<sup>b</sup> |
| --- | --- | --- | --- | --- | --- | --- | --- | --- | --- | --- |
| A2–Asthma | rs34712979 | 4:106819053 | A | −0.102 | 0.047 | $5.5 \times 10^{-10}$ | $3.3 \times 10^{-18}$ | ↑↓ | *NPNT* | Resp |
| | rs34517439 | 1:78450517 | A | 0.11 | 0.034 | $1.0 \times 10^{-6}$ | $3.6 \times 10^{-6}$ | ↑↑ | *DNAJB4; GIPC2* | Resp, Imm |
| A2–ILD | rs12802931 | 11:1236164 | G | −0.103 | 0.583 | $2.4 \times 10^{-8}$ | $6.8 \times 10^{-45}$ | ↑↓ | *MUC5B* | — |

| Comparison | Lead SNP | Chr:Pos | EA | $\beta$_COVID | $\beta$_Resp | P_COVID | P_Resp | Dir.<sup>a</sup> | Nearest gene(s) | PheWAS<sup>b</sup> |
| --- | --- | --- | --- | --- | --- | --- | --- | --- | --- | --- |
| | rs2076295 | 6:7563232 | G | 0.068 | 0.155 | $1.5 \times 10^{-7}$ | $7.7 \times 10^{-10}$ | ↑↑ | *DSP* | Resp |
| A2–IPF | rs12585036 | 13:113535741 | T | 0.138 | −0.121 | $9.3 \times 10^{-19}$ | $2.4 \times 10^{-11}$ | ↑↓ | *ATP11A* | Resp, Imm |
| | rs2277732 | 19:4723670 | A | 0.237 | 0.102 | $2.4 \times 10^{-56}$ | $3.6 \times 10^{-9}$ | ↑↑ | *DPP9* | — |
| | rs12802931 | 11:1236164 | G | −0.103 | 0.53 | $2.4 \times 10^{-8}$ | $9.6 \times 10^{-112}$ | ↑↓ | *MUC5B* | — |
| | rs2076295 | 6:7563232 | G | 0.068 | 0.192 | $1.5 \times 10^{-7}$ | $3.5 \times 10^{-40}$ | ↑↑ | *DSP* | Resp |
| B2–Asthma | rs34712979 | 4:106819053 | A | −0.061 | 0.047 | $3.8 \times 10^{-8}$ | $3.3 \times 10^{-18}$ | ↑↓ | *NPNT* | Resp |
| | rs13135092 | 4:103198082 | G | 0.085 | 0.067 | $2.7 \times 10^{-7}$ | $5.6 \times 10^{-11}$ | ↑↑ | *SLC39A8* | Resp, Imm |
| B2–ILD | rs12802931 | 11:1236164 | G | −0.067 | 0.583 | $9.9 \times 10^{-8}$ | $6.8 \times 10^{-45}$ | ↑↓ | *MUC5B* | — |
| B2–IPF | rs75898026 | 13:113561082 | A | 0.086 | −0.111 | $9.4 \times 10^{-14}$ | $3.2 \times 10^{-9}$ | ↑↓ | *MCF2L* | Resp, Imm |
| | rs12802931 | 11:1236164 | G | −0.067 | 0.53 | $9.9 \times 10^{-8}$ | $9.6 \times 10^{-112}$ | ↑↓ | *MUC5B* | — |
| | rs35574495 | 19:4686988 | T | 0.125 | 0.104 | $8.7 \times 10^{-23}$ | $1.8 \times 10^{-6}$ | ↑↑ | *DPP9* | — |
| C2–Asthma | rs13135092 | 4:103198082 | G | 0.046 | 0.067 | $2.5 \times 10^{-8}$ | $5.6 \times 10^{-11}$ | ↑↑ | *SLC39A8* | Resp, Imm |
| C2–ILD | rs13243708 | 7:99616291 | C | 0.025 | 0.138 | $4.4 \times 10^{-7}$ | $6.2 \times 10^{-8}$ | ↑↑ | *ZKSCAN1* | Imm |
| C2–IPF | rs732631 | 19:4719025 | G | 0.033 | 0.093 | $3.9 \times 10^{-12}$ | $4.1 \times 10^{-9}$ | ↑↑ | *DPP9* | — |

<sup>a</sup> ↑↑, concordant direction (same allele increases risk for both traits); ↑↓, discordant direction (risk allele for one trait is protective for the other).
<sup>b</sup> Lead SNP or proxy previously associated (Bonferroni-corrected $P < 0.05$ ) with respiratory (Resp) and/or immunological (Imm) phenotypes in GWAS Atlas PheWAS. — indicates no significant association in these domains.
EA, effect allele. All loci validated by three independent methods (cofdr, gwas-pw, HyPrColoc). Genome-wide likelihood ratio tests confirmed significant shared genetic architecture for all comparisons shown (all LRT. $P \approx 0$ ; see Supplementary Table 2.2).

The comparison between A2 and idiopathic pulmonary fibrosis (IPF) yielded the highest number of validated risk loci across the 24 pairwise analyses (Table 2). The four-group cofdr model estimated that A2 and IPF share approximately 3.31% of associated SNPs. Furthermore, the likelihood ratio test provided strong evidence for a shared genetic basis between these two traits (*LRT*. *p* ≈ 0) (Supplementary Table 2.2).

PheWAS via GWAS Atlas revealed that five shared SNPs were significantly associated with other respiratory or immunological traits. Three SNPs (rs34517439 for A2-Asthma, rs13135092 for B2-Asthma, and rs13135092 for C2-Asthma) demonstrated consistent allelic directions, while two (rs12585036 for A2-IPF and rs75898026 for B2-IPF) exhibited opposing effects (Table 3).

#### Pairwise shared signals at the gene level

By integrating GWAS data with multi-tissue transcriptome and proteome data, we identified shared genes using gene-based cofdr. We prioritized genes that were consistently identified by at least three of our four analytical pipelines: MAGMA-cofdr, TWAS-cofdr, spTWAS-cofdr, and PWAS-cofdr. This approach identified 67 validated shared genes for A2 and related respiratory disorders, 55 for B2, and 26 for C2 (Table 2). Of these, 65 genes reached FDR<0.05 in the original MAGMA analysis for both traits. In the original TWAS analysis for lung and whole blood tissues, 23 genes were significant for both traits, with 5 showing concordant effect directions. Splicing-level analysis via spTWAS identified 59 genes significant in both traits, with 20 showing concordant directions. PWAS identified 3 shared proteins, one of which exhibited a consistent effect direction (Table 4).

**Table 4.** Key shared genes with multi-omics convergent evidence.

| Gene | Comparison<sup>a</sup> | MAGMA $\pi$_TT | TWAS $\pi$_TT (tissue) | spTWAS $\pi$_TT (tissue) | PWAS $\pi$_TT | Dir.<sup>b</sup> | Functional annotation |
| --- | --- | --- | --- | --- | --- | --- | --- |
| **COVID–IPF/ILD shared signals** |  |  |  |  |  |  |  |
| *ATP11A* | A2–IPF | 0.99 | 0.97 (Blood) | 1.00 (Lung) | — | ↑↓ | Phospholipid flippase; lung fibrosis |
| *DPP9* | A2–IPF | 1 | — | 0.99 (Lung) | — | ↑↑ | Dipeptidyl peptidase; known COVID–IPF locus |
| *TRIM4* | A2–IPF | 0.96 | — | 0.99 (Lung) | — | ↑↑ | E3 ubiquitin ligase; IFN-β induction |
| *BAG6* | A2–IPF | 0.99 | — | 0.98 (Lung) | — | ↑↑ | Co-chaperone; NK cell regulation |
| *ZNF3* | B2–IPF | 1 | 0.93(Blood) | 1.00 (Lung) | — | ↑↑ | Zinc-finger transcription factor |
| *ZSCAN21* | B2–IPF | 1 | — | 1.00 (Lung) | — | ↑↑ | Zinc-finger transcription factor |
| *FUT3* | C2–IPF | 0.94 | — | — | 0.99 | ↑↑ | Fucosyltransferase; glycan biology |
| **COVID–Asthma shared signals** |  |  |  |  |  |  |  |
| *TCF19* | A2–Asthma | 1 | 1.00 (Blood) | 1.00 (Lung) | — | ↑↓ | Transcription factor; MHC region |
| *MED24* | A2–Asthma | 0.96 | 0.96 (Blood) | 1.00 (Lung) | — | ↑↓ | Mediator complex; transcriptional control |
| *INTS12* | A2–Asthma | 0.99 | 0.99 (Blood) | 1.00 (Lung) | — | ↑↓ | Integrator complex; snRNA processing |
| *NCOR1* | A2–Asthma | 0.94 | 0.96 (Blood) | 0.98 (Lung) | — | ↑↓ | Nuclear co-repressor; macrophage inflammation |
| *ZSWIM7* | B2–Asthma | 0.99 | 0.97 (Lung) | 0.99 (Lung) | — | ↑↑ | DNA repair; homologous recombination |
| *NPNT* | A2–Asthma | — | — | 1.00 (Lung) | 1 | ↑↓ | Nephronectin; integrin signaling |
| *ICAM1* | B2–Asthma | 0.96 | 0.98 (Blood) | — | — | ↑↑ | Cell adhesion; leukocyte migration |
| *BPTF* | A2–Asthma | 0.93 | — | 0.93 (Lung) | — | ↑↑ | Chromatin remodeling (NURF complex) |
| *ACSL6* | B2–Asthma | 0.99 | 0.91 (Blood) | — | — | ↑↑ | Long-chain fatty acid metabolism |
| *PIGL* | B2–Asthma | 0.95 | — | 0.96 (Blood) | — | ↑↑ | GPI anchor biosynthesis |
| *GSDMA* | C2–Asthma<sup>c</sup> | 0.99 | 1.00 (Lung) | — | — | ↑↓ | Gasdermin A; pyroptosis |
| *MUC1* | C2–Asthma | 1 | — | 1.00 (Lung) | — | ↑↑ | Mucin 1; mucosal barrier defense |
| *CCHCR1* | A2–Asthma | 1 | 0.90 (Lung) | 1.00 (Blood) | — | ↑↓ | HLA region; immune regulation |
| *PSMD3* | C2–Asthma | 1 | — | — | — | — | Proteasome subunit; protein degradation |
| *MICB* | A2–Asthma | 0.91 | — | — | — | — | MHC class I; NK cell activation |
| **Other notable shared signals** |  |  |  |  |  |  |  |
| *SURF1* | C2–COPD | 0.93 | — | 0.99 (Blood) | — | ↑↑ | Cytochrome c oxidase assembly |
| *DEPTOR* | B2–IPF | 1 | 0.99 (Lung) | — | — | ↑↑ | mTOR inhibitor; autophagy regulation |
| *GSDMB* | B2–COPD | 1 | 1.00 (Lung) | 1.00 (Lung) | — | ↑↓ | Gasdermin B; airway inflammation |

<sup>a</sup> The comparison with the strongest evidence is shown. Many genes appear across multiple comparisons (see Supplementary Table 2.3-2.6 for complete results). For example, *GSDMA* was identified in A2–Asthma (0.95), B2–Asthma (0.99), A2–COPD (0.97), B2–COPD (0.98), C2–Asthma (0.99), and C2–COPD (0.99); *MED24* was identified across all three COVID phenotypes with both asthma and COPD.
<sup>b</sup> ↑↑, concordant direction; ↑↓, discordant (opposing) direction.
<sup>c</sup> $\pi$ \_TT is shown for C2–Asthma; also shared in five additional comparisons (see footnote a).
Dashes (—) indicate the gene was not identified at the specified omics layer at the threshold used ( $\pi$ \_TT ≥ 0.9 for MAGMA/TWAS/spTWAS; ≥ 0.8 for PWAS). TWAS and spTWAS analyses were conducted across 49 tissues; for clarity, only results from lung and whole blood are shown here. Full tissue-specific results are available in Supplementary Tables 2.4–2.5.

The A2-IPF comparison again yielded the most validated shared genes (Table 2). Notably, TWAS-cofdr identified *ATP11A* in whole blood with a high posterior probability (π_TT = 0.97). Interestingly, the effect direction was discordant: increased expression of *ATP11A* in blood was associated with higher IPF risk but was protective against severe COVID-19 (A2). This gene was also prioritized by spTWAS-cofdr with high posterior probabilities across tissues (π_TT = 1.00 in lung; π_TT = 0.99 in blood) and by MAGMA-cofdr (π_TT = 0.99) (Table 4).

PheWAS indicated that most identified genes were associated with respiratory or immunological traits, with the exceptions of *UNC50* and *ABO*. Pathway enrichment via FUMA highlighted immunoregulatory interactions between lymphoid and non-lymphoid cells for A2-Asthma, lung function (FEV1) for A2-Asthma and B2-Asthma, and white blood cell counts for C2-COPD (Supplementary Data 2).

#### Multiple-trait shared signals at the SNP and gene levels

Using a posterior probability threshold of 0.9, cofdr identified ten loci shared across A2 and four respiratory disorders (asthma, ILD, IPF, and COPD); however, these were not detected by hyprcoloc. A more permissive threshold of 0.8 yielded eight loci for B2 and two for C2 (Table 2). At the gene level, MAGMA-cofdr and TWAS-cofdr identified two and three shared genes, respectively (Supplementary Table 3.3-3.4).

### Results of differentiation analyses

#### Pairwise differentiation analyses at the SNP level

Differential association tests across the 24 pairwise comparisons were conducted using DDx and validated with CC-GWAS. We identified 69, 58, and 87 validated differentiated loci for A2, B2, and C2, respectively (Table 2). Using a significance threshold of *P* < 5 × 10^−8^, these loci were categorized into four groups: (1) 80 loci significant only in COVID-19; (2) 118 significant only in other respiratory disorders; (3) 7 significant in both traits but with greater significance in COVID-19 (all showing opposing effects except rs2277732 in A2 vs. IPF); and (4) 9 loci not significant for either trait individually, which could be reclassified as unique or opposing-direction loci using a more lenient threshold of p < 5 × 10^−6^. Particularly, PheWAS of those 80 COVID-19-unique loci revealed that 21 lead SNPs were previously associated with other respiratory or immunological studies, mostly with opposing effects (Supplementary Table 4.2).

#### Pairwise differentiation analyses at the gene level

We merged results from four gene-level differentiation pipelines: MAGMA-DDx/CC-GWAS, TWAS-DDx/CC-GWAS, spTWAS-DDx/CC-GWAS, and PWAS-DDx/CC-GWAS. Genes identified by at least three approaches were considered validated, resulting in 95, 90, and 183 genes with differential expression/splicing/protein levels for A2, B2, and C2 comparisons, respectively (Table 2).

Among all validated genes, 99 had FDR < 0.05 in the original MAGMA gene-level analysis for COVID-19 but FDR > 0.05 for the respiratory comparator; 27 had FDR < 0.05 in the original TWAS analysis in lung and whole blood for COVID-19 but FDR > 0.05 for the comparator, all with opposing effect directions; 94 had FDR < 0.05 in the original spTWAS analysis in lung and whole blood for COVID-19 but FDR > 0.05 for the comparator, of which 72 showed opposing effects; and 3 had FDR < 0.05 in the original PWAS analysis for COVID-19 but FDR > 0.05 for the comparator, all with opposing effects (Table 5; Supplementary Table 4.3-4.6).

**Table 5.** Key differential genes distinguishing COVID-19 from other respiratory disorders.

| Gene | Comparison<sup>a</sup> | Best DDx FDR | FDR_COVID | FDR_Resp | Omics<sup>b</sup> | Dir.<sup>c</sup> | Functional theme |
| --- | --- | --- | --- | --- | --- | --- | --- |
| **Interferon signaling** |  |  |  |  |  |  |  |
| *IFNAR2* | A2 vs. COPD | sT: 1.8×10<sup>−31</sup> | 3.0×10<sup>−36</sup> | 0.86 | M, sT | COVID ↑ | Type I IFN receptor |
|  | A2 vs. IPF | sT: 6.7×10<sup>−13</sup> | 3.0×10<sup>−36</sup> | 0.92 | M, sT | COVID ↑ |  |
| *OAS1* | A2 vs. IPF | sT: 4.0×10<sup>−5</sup> | 6.8×10<sup>−8</sup> | 0.74 | M, sT | COVID ↑ | 2'-5' Oligoadenylate synthetase |
| *OAS3* | A2 vs. IPF | T: 9.3×10<sup>−3</sup> | 5.3×10<sup>−5</sup> | 0.58 | M, T | COVID ↑ | 2'-5' Oligoadenylate synthetase |
| *TYK2* | A2 vs. IPF | sT: $2.1 \times 10^{-7}$ | $2.5 \times 10^{-8}$ | 0.44 | M, sT | COVID ↑ | JAK–STAT signaling kinase |
| *IL10RB* | A2 vs. COPD | M: $1.6 \times 10^{-5}$ | $1.4 \times 10^{-5}$ | 0.41 | M | COVID ↑ | IL-10 receptor; anti-inflammatory |
| ***Vascular / endothelial biology*** |  |  |  |  |  |  |  |
| *ARHGAP27* | A2 vs. Pneumonia | M: $3.4 \times 10^{-7}$ | $9.1 \times 10^{-7}$ | 0.88 | M, T, sT | Opposite | Rho GTPase; endothelial barrier |
| *RASIP1* | A2 vs. Pneumonia | M: $1.9 \times 10^{-6}$ | $5.9 \times 10^{-6}$ | 0.61 | M, T, sT | Opposite | Endothelial junction assembly |
| ***Extracellular matrix / tissue remodeling*** |  |  |  |  |  |  |  |
| *THBS3* | A2 vs. COPD | M: $1.6 \times 10^{-11}$ | $1.1 \times 10^{-10}$ | 0.16 | M, sT | COVID ↑ | Thrombospondin; ECM |
| *MTX1* | A2 vs. COPD | sT: $7.7 \times 10^{-11}$ | $7.9 \times 10^{-10}$ | 0.3 | M, sT | Opposite | Mitochondrial import; organellar stress |
| ***Autophagy / intracellular trafficking*** |  |  |  |  |  |  |  |
| *FYCO1* | A2 vs. IPF | M: $7.3 \times 10^{-9}$ | $3.7 \times 10^{-7}$ | 0.82 | M, sT | COVID ↑ | Autophagosome transport |
| *FUT2* | A2 vs. IPF | M: $5.2 \times 10^{-5}$ | $2.5 \times 10^{-7}$ | 0.63 | M, T | COVID ↑ | Fucosyltransferase; mucosal immunity |
| *CALCOCO2* | B2 vs. Asthma | T: $3.9 \times 10^{-4}$ | $1.5 \times 10^{-2}$ | 0.06 | T | Opposite | Selective autophagy receptor (xenophagy) |
| *PLEKHM1* | A2 vs. Pneumonia | M: $3.6 \times 10^{-7}$ | $4.1 \times 10^{-6}$ | 0.88 | M, T, sT | Opposite | Autophagy–lysosomal trafficking |
| ***NF-κB / inflammatory signaling*** |  |  |  |  |  |  |  |
| *NFKBIZ* | C2 vs. Pneumonia | M: $5.4 \times 10^{-10}$ | $3.0 \times 10^{-9}$ | 0.27 | M | COVID ↑ | NF-κB inhibitor zeta; innate immunity |
| *FBRSL1* | A2 vs. Asthma | M: $1.5 \times 10^{-6}$ | $4.6 \times 10^{-6}$ | 0.14 | M, T, sT | COVID ↑ | Fibrosin-like 1; chromatin regulation |
| *MAMSTR* | A2 vs. Pneumonia | M: $9.8 \times 10^{-7}$ | $1.8 \times 10^{-5}$ | 0.54 | M | COVID ↑ | MEF2-activating; transcriptional regulation |
| ***Other notable differential signals*** |  |  |  |  |  |  |  |
| *DPP9* | A2 vs. Pneumonia | M: $9.9 \times 10^{-10}$ | $5.6 \times 10^{-10}$ | 1 | M, T, sT | COVID ↑ | Dipeptidyl peptidase |
| *LTF* | A2 vs. Asthma | M: $3.3 \times 10^{-7}$ | $1.2 \times 10^{-6}$ | 0.16 | M | COVID ↑ | Lactoferrin; innate immunity |
| *GSDMB* | B2 vs. Pneumonia | sT: $2.7 \times 10^{-4}$ | $1.0 \times 10^{-3}$ | 0.6 | M, T, sT | Opposite | Gasdermin B; pyroptosis |
| *ABO* | C2 vs. Pneumonia | P: $7.3 \times 10^{-12}$ | $2.5 \times 10^{-12}$ | 0.9 | M, T, P | COVID ↑ | Blood group; coagulation |

<sup>a</sup> The comparison with the strongest or most illustrative result is shown; many genes are differential across multiple comparisons (see Supplementary Table 4.3–4.6 for complete results).
<sup>b</sup> Omics layers in which the gene was identified as differential: M = MAGMA, T = TWAS, sT = spTWAS, P = PWAS. TWAS and spTWAS were conducted across 49 tissues; only lung and whole blood results are shown here. Full results are provided in Supplementary Tables 4.4–4.5.
<sup>c</sup> "COVID ↑" = significant in COVID-19 only (FDR < 0.01) but not in the respiratory comparator (FDR > 0.05). "Opposite" = significant in COVID-19 but with opposing effect directions between COVID-19 and the comparator.
FDR\_COVID and FDR\_Resp refer to the original (non-differential) GWAS, demonstrating that these genes are driven primarily by COVID-19 association.

PheWAS analysis demonstrated that most differential genes had been linked to measures of pulmonary function and/or immune response. Biological processes associated with differential GWAS were further explored using MAGMA gene-set analysis; we particularly focused on gene sets significant for COVID-19 only (FDR < 0.05) (Table 7 Panel A; Supplementary Table 4.7). We highlight some of the pathways below. The “DEFECTIVE_CSF2RB_CAUSES_SMDP5” pathway, identified in 11 of 24 pairwise differential analyses, highlights the distinct role of alveolar macrophages in COVID-19. The “RESPONSE_TO_TYPE_III_INTERFERON” gene set was detected in 6 A2-related analyses (A2 vs. asthma/COPD/pneumonia(all)/bacterial pneumonia/viral pneumonia/influenza), indicating its role in distinguishing severe COVID-19 from other respiratory conditions. The A2 vs. asthma and B2 vs. asthma comparisons identified the gene set “TYPE_I_INTERFERON_INDUCTION_AND_SIGNALING” as unique to severe and hospitalized COVID-19, whereas C2 vs. asthma revealed enrichment of “NEGATIVE_REGULATION_OF_TYPE_II_INTERFERON_PRODUCTION.“

Besides, we also performed an exploratory drug enrichment (over-representation) analysis using WebGestalt and EnrichR (Supplementary Table 4.10-4.11), based on the significant COVID-specific genes from MAGMA-DDx and TWAS-DDx identified in the comparisons of COVID-19 vs flu/pneumonia (as extracted from Supplementary Table 4.3-4.4). For example, some top enriched drugs included interferons, MMR vaccine, and ribavirin. These drugs were supported by several previous studies and may improve COVID-19 outcomes^62–65^, although such evidence remains tentative and requires further studies.

Tissue and cell-type specificity analyses were also conducted using FUMA (Table 7 Panel B; Supplementary Table 4.8-4.9). The involvement of lung and spleen tissue was demonstrated as specific to hospitalized COVID-19 (B2) when compared to (bacterial/viral) pneumonia or influenza. No cell types were identified as uniquely enriched for COVID-19.

### Multi-trait differentiation at the SNP and gene levels

In mtCOJO analysis, the number of genome-wide significant SNPs decreased after conditioning, consistent with pleiotropy between disorders. However, 20 of the 2,702 significant SNPs from the unadjusted severe COVID-19 (A2) became more significant after conditioning, forming a risk locus near *MAPT*. Similarly, 5 SNPs in the hospitalized COVID-19 (B2) showed increased significance after conditioning, forming a locus near *CCR3* (Table 6 Panel A). Conversely, all SNPs in the reported COVID-19 (C2) showed decreased significance, indicating broad pleiotropy.

**Table 6.** COVID-19-specific signals from multi-trait conditional analysis (mtCOJO)

**Panel A. Lead SNPs with increased significance after conditioning on respiratory traits**
| COVID phenotype | Lead SNP | Chr:Position | Nearest gene(s) | β_cond | P_cond | β_unadj | P_unadj | PheWAS |
| --- | --- | --- | --- | --- | --- | --- | --- | --- |
| A2 (severe) | rs17652337 | 17:44083323 | *MAPT* | −0.20 | $4.7 \times 10^{-12}$ | −0.12 | $1.2 \times 10^{-11}$ | Resp, Imm |
| B2 (hospitalized) | rs1491957 | 3:46274960 | *CCR3* | 0.1 | $3.0 \times 10^{-11}$ | 0.05 | $3.8 \times 10^{-10}$ | Imm |

**Panel B. Validated COVID-specific genes across omics layers**
| COVID phenotype | Gene | Layer | Tissue | Z_cond | FDR_cond | Z_unadj | FDR_unadj | Stronger?<sup>a</sup> | PheWAS |
| --- | --- | --- | --- | --- | --- | --- | --- | --- | --- |
| A2 | *FYCO1* | MAGMA | — | — | $3.6 \times 10^{-8}$ | — | $3.7 \times 10^{-7}$ | ✓ | Imm |
| | | spTWAS | Lung | 4.85 | $1.9 \times 10^{-3}$ | 7.35 | $5.0 \times 10^{-10}$ | — | |
| | *HCN3* | MAGMA | — | — | $4.3 \times 10^{-3}$ | — | $5.1 \times 10^{-3}$ | ✓ | Resp, Imm |
| | | TWAS | Lung | -4.87 | $4.3 \times 10^{-3}$ | -4.23 | $1.1 \times 10^{-2}$ | ✓ | |
| | | TWAS | Blood | -4.87 | $1.9 \times 10^{-3}$ | -4.23 | $1.0 \times 10^{-2}$ | ✓ | |
| | *IFNAR2* | MAGMA | — | — | $1.5 \times 10^{-6}$ | — | $3.7 \times 10^{-7}$ | — | — |
| | | spTWAS | Lung | 8.63 | $6.6 \times 10^{-14}$ | 13.26 | $3.0 \times 10^{-36}$ | — | |
| | *ICAM5* | MAGMA | — | — | $8.6 \times 10^{-4}$ | — | $5.2 \times 10^{-10}$ | — | Resp, Imm |
| | | PWAS | — | 5.03 | $6.1 \times 10^{-4}$ | -5.60 | $2.7 \times 10^{-5}$ | ✓ | |
| B2 | *NXPE3* | MAGMA | — | — | $1.3 \times 10^{-3}$ | — | $1.6 \times 10^{-8}$ | — | Imm |
| | *OAS1* | MAGMA | — | — | $5.5 \times 10^{-3}$ | — | $6.9 \times 10^{-10}$ | — | Imm |
| | | spTWAS | Lung | 4.69 | $3.5 \times 10^{-3}$ | 7.47 | $3.0 \times 10^{-10}$ | — | |
| C2 | *NXPE3* | MAGMA | — | — | $8.1 \times 10^{-6}$ | — | $4.5 \times 10^{-7}$ | — | Imm |
| | *ZBTB11* | MAGMA | — | — | $1.4 \times 10^{-5}$ | — | $4.5 \times 10^{-7}$ | — | Imm |

<sup>a</sup>✓ indicates the gene's association with COVID-19 became more significant after conditioning on all genetically correlated respiratory traits, suggesting COVID-19 specificity independent of shared respiratory genetics. Dashes indicate the association remained significant but did not increase.

Genes detected by at least three of the four approaches (MAGMA-mtCOJO, TWAS-mtCOJO, spTWAS-mtCOJO, and PWAS-mtCOJO) were considered prioritized/validated genes associated with the conditioned traits. The conditional analyses of A2, B2, and C2 revealed 4, 2, and 2 validated differential genes, respectively (Table 2).

Among four significant associations of severe COVID-19 detected by MAGMA-mtCOJO, *FYCO1* and *HCN3* showed greater significance in the conditional analysis compared to the unadjusted analysis (Table 6 Panel B). Further TWAS-mtCOJO analysis shows that lower genetically predicted expression of *HCN3* in lung and whole blood tissues was associated with increased risk of severe COVID-19. Specifically, this association strengthened in the conditional A2 analysis. These findings suggest that *FYCO1* and *HCN3* may be uniquely linked to severe COVID-19 rather than general respiratory diseases. Finally, MAGMA gene-set and tissue-property analyses identified two gene sets and three tissues that only reached significance after conditioning, further refining the COVID-19-specific genetic signature (Supplementary Table 5.7-5.8; Table 7 Panel B).

**Table 7.** COVID-discriminating pathways and tissue enrichments.

| Pathway / Gene set | Source | N<sup>a</sup> | COVID pheno. | Best FDR_DDx (comparison) | FDR_COVID<sup>b</sup> | FDR_Resp<sup>b</sup> | Biological theme |
| --- | --- | --- | --- | --- | --- | --- | --- |
| *Type I / general interferon signaling* |  |  |  |  |  |  |  |
| Overview of IFN-mediated signaling | WP | 4 | A2 | $3.4 \times 10^{-5}$ (A2 vs. Pneu.) | $7.6 \times 10^{-4}$ | 0.95 | Type I IFN |
| Regulation of IFNA/IFNB signaling | REACTOME | 5 | A2, B2 | $2.9 \times 10^{-4}$ (A2 vs. Pneu.) | $2.7 \times 10^{-3}$ | 0.88 | Type I IFN regulation |
| Interferon alpha/beta signaling | REACTOME | 4 | A2 | $2.6 \times 10^{-3}$ (A2 vs. Pneu.) | 0.023 | 0.95 | Type I IFN |
| IFNA pathway | BIOCARTA | 2 | A2 | $6.5 \times 10^{-3}$ (A2 vs. Bact. pneu.) | 0.023 | 0.89 | Type I IFN |
| IFN-mediated signaling pathway | GOBP | 2 | A2 | $3.7 \times 10^{-2}$ (A2 vs. Bact. pneu.) | 0.095 | 0.99 | IFN (general) |
| Interferon receptor activity | GOMF | 2 | A2 | $2.1 \times 10^{-2}$ (A2 vs. Pneu.) | 0.037 | 0.98 | IFN receptor |
| *Type III interferon signaling* |  |  |  |  |  |  |  |
| Response to type III interferon | GOBP | 4<sup>c</sup> | A2 | $4.7 \times 10^{-3}$ (A2 vs. Bact. pneu.) | 0.014 | 0.95–0.97 | Type III IFN |
| Type III interferon signaling | WP | 2 | A2 | 1.5×10<sup>-2</sup> (A2 vs. Bact. pneu.) | 0.037 | 0.86–0.99 | Type III IFN |
| ***GM-CSF / alveolar macrophage / surfactant*** |  |  |  |  |  |  |  |
| Defective CSF2RB causes SMDP5 | REACTOME | 9<sup>d</sup> | A2, B2, C2 | 1.4×10<sup>-5</sup> (B2 vs. Flu) | 2.8×10<sup>-5</sup> | 0.69–0.97 | GM-CSF / macrophage |
| Diseases assoc. with surfactant metabolism | REACTOME | 12 | A2, B2, C2 | 1.7×10<sup>-5</sup> (B2 vs. Flu) | 2.7×10<sup>-4</sup> | 0.44–0.98 | Surfactant dysfunction |
| Surfactant homeostasis | GOBP | 3 | A2 | 1.9×10<sup>-2</sup> (A2 vs. Viral pneu.) | 0.021 | 0.74–1.00 | Type II AEC |
| Alveolar lamellar body | GOCC | 1 | A2 | 3.8×10<sup>-2</sup> (A2 vs. Viral pneu.) | 0.065 | 1 | AEC organelle |
| ***Innate immune / antimicrobial response*** |  |  |  |  |  |  |  |
| Immune response to tuberculosis | WP | 7 | A2, B2 | 1.1×10<sup>-3</sup> (A2 vs. Bact. pneu.) | 7.0×10<sup>-4</sup> | 0.92–0.99 | Innate immunity |
| Eosinophils pathway | BIOCARTA | 2 | B2 | 2.4×10<sup>-2</sup> (B2 vs. Viral pneu.) | 5.4×10<sup>-6</sup> | 0.88–0.97 | Eosinophil biology |
| Measles virus infection | WP | 1 | B2 | 3.3×10<sup>-2</sup> (B2 vs. Viral pneu.) | 0.024 | 0.94 | Antiviral response |
| ***Other notable COVID-specific pathways*** |  |  |  |  |  |  |  |
| Multivesicular body lumen | GOCC | 3 | A2 | 3.3×10<sup>-2</sup> (A2 vs. Flu) | 0.082 | 0.83–0.99 | Endosomal trafficking |
| Lung fibrosis | WP | 2 | A2 | 1.9×10<sup>-2</sup> (A2 vs. Viral pneu.) | 0.24 | 0.95–0.97 | Fibrotic remodeling |
| 1q21 amplicon (breast cancer) | NIKOLSKY | 12 | A2, B2, C2 | 1.1×10<sup>-5</sup> (A2 vs. Flu) | 6.5×10<sup>-6</sup> | 0.84–0.99 | Epidermal barrier<sup>e</sup> |

<sup>a</sup> Number of pairwise COVID vs. respiratory comparisons in which the gene set reached FDR\_DDx < 0.05 while FDR\_Resp > 0.05 (non-significant in the respiratory comparator).
<sup>b</sup> FDR\_COVID and FDR\_Resp shown for the comparison with the best FDR\_DDx. FDR\_COVID reflects significance in the original COVID-19 GWAS; FDR\_Resp reflects significance in the original respiratory comparator GWAS.
<sup>c</sup> In the provided supplementary data; the manuscript text reports this pathway in 6 A2 comparisons (including A2 vs. Asthma and A2 vs. COPD, which may appear in separate supplementary files).
<sup>d</sup> 9 comparisons present in the provided supplementary data; the manuscript text reports 11 of 24 comparisons.
<sup>e</sup> The 1q21 amplicon gene set contains *LCE3* cluster genes involved in epidermal differentiation and barrier function; these genes have also been associated with psoriasis and atopic dermatitis, suggesting a COVID-specific epithelial barrier disruption pathway.

Panel B. Tissues enriched specifically in COVID-19 differential or conditional GWAS (MAGMA gene-property analysis)
| Tissue | Analysis | Comparison(s) | COVID phenotype | Sig. in unadj. COVID GWAS? | Sig. in resp. GWAS? |
| --- | --- | --- | --- | --- | --- |
| Lung | DDx | B2 vs. Pneumonia, B2 vs. Bact. pneu., B2 vs. Influenza | B2 | Yes | No |
| Spleen | DDx | B2 vs. Pneumonia, B2 vs. Bact. pneu., B2 vs. Viral pneu., B2 vs. Influenza | B2 | **No** | No |
| Lung | mtCOJO | A2 conditioned | A2 | Yes | — |
| Spleen | mtCOJO | A2 conditioned | A2 | **No** | — |
| Thyroid | mtCOJO | A2 conditioned | A2 | **No** | — |

Bold **No** highlights tissues that emerge as significant only after differential or conditional analysis, i.e., they were masked in the original unadjusted GWAS and represent newly identified COVID-discriminating tissue signals. The spleen finding is consistent with clinical reports of splenic white pulp depletion and immune cell apoptosis in COVID-19 patients (see main text).

## Discussion

### Overview

Here we disentangled shared and disease-discriminating (differential) genetic contributions between COVID-19 outcomes (A2/B2/C2) and eight respiratory risk factors/disorders using a unified summary-statistics framework across 24 pairwise and three multi-trait comparisons, at a multi-omics level (Figure 1). Rather than evaluating “genetic overlap” per se, we explicitly decomposed the genetic architecture into: (i) shared association signals (variants/genes affecting multiple traits) and (ii) differential signals (variants/genes showing heterogeneous effects between COVID-19 and related respiratory phenotypes). By integrating SNP-level association patterns with multi-omics gene-level evidence (MAGMA, TWAS, spTWAS, PWAS), we moved from the SNP level to mechanistically interpretable hypotheses about various molecular layers, including expression, splicing, and protein abundance, through which genetic risk may act.

**Figure 1.**
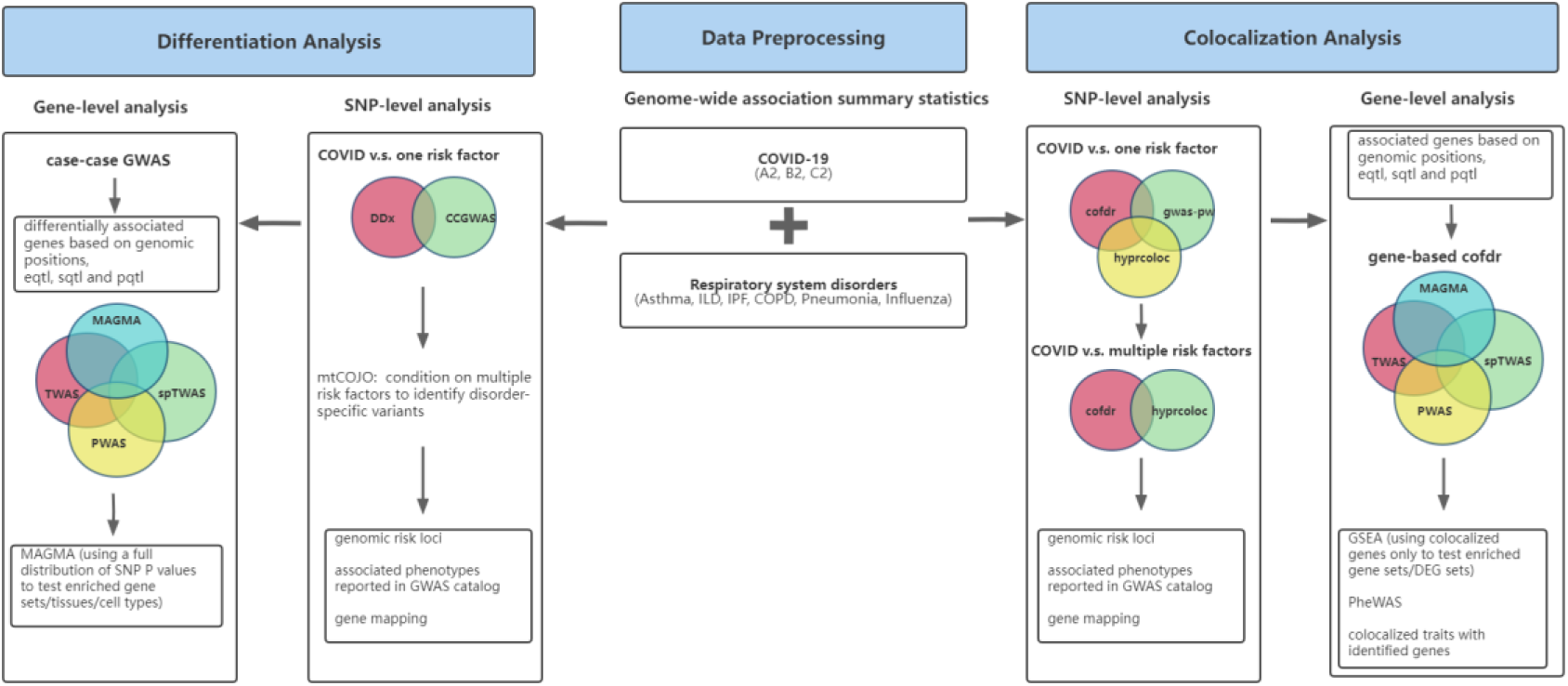
The framework of colocalization and differentiation analyses.

This work complements but is distinctly different from other studies focused on revealing genetic factors for individual respiratory infections or disorders. The current study, for example, is fundamentally different from our recent GWAS on hospitalized influenza^38^ which focused on discovering specific influenza risk loci (e.g., *ST6GAL1*) and genetic factors underlying time-to-hospitalization. The present study treats influenza gwas statistics as just one component of a broader analysis, where COVID-19 serves as the index trait. In addition., whereas the influenza study only included a single pairwise comparison, the current work encompasses 24 pairwise and 3 multi-trait comparisons involving three COVID-19 phenotypes (A2/B2/C2; only B2 was covered in ref^38^) and a wide spectrum of eight respiratory disorders. We employed multiple complementary methods for both shared (cofdr, gwas-pw, hyprcoloc) and differential (DDx, CC-GWAS, mtCOJO) signal detection and systematically integrate multi-omics layers. The multi-trait conditional analysis (mtCOJO), which isolates COVID-specific signals, is also unique to this study. Here, our objective is not to characterize individual respiratory infections as in ref^38^ or other GWAS, but rather to apply a unified framework to systematically disentangle the shared “pan-respiratory” genetic backbone from the specific molecular mechanisms (e.g. splicing defects, interferon dysregulation) that distinguish COVID-19 from the wider spectrum of respiratory diseases.

### Novelty of the proposed analytic framework

Our study provides a new, coherent, and comprehensive analytic framework for translating large-scale GWAS summary statistics into interpretable shared-versus-differential genetic insights across many traits. This framework is potentially applicable to other complex diseases beyond COVID-19 (Figure 1).

Prior work on shared genetics often focused on global genetic correlation, overlap of significant loci, or cross-trait meta-analysis. While these approaches capture comorbidity, they typically do not reveal trait-discriminating biology. Here, we combined complementary tools to answer two distinct questions: *Which genetic signals are shared?* and *Which signals differentiate COVID-19 from other related respiratory conditions?* This “two-axis” architecture (shared vs. differential) is central to generating biological insights and providing a comprehensive picture of the genetic architecture.

Furthermore, the above-mentioned methods also have limitations in studying the genetics of disease comorbidities. Global genetic correlation is commonly performed, but this method cannot discern individual loci shared among disorders. The study of overlapping significant loci is straightforward, but it cannot provide a quantitative measure of the probability of shared signals, and moderate-effect SNPs/genes may often be missed. Cross-trait meta-analysis is also a common strategy, but it does not provide a direct measure of the probability that a variant or gene is shared across phenotypes. Instead, it provides a measure of the significance of aggregate association across traits. Our proposed framework using cofdr (a flexible algorithm which detects shared/pleiotropic genetic signals) and coloc-type algorithms (uncovering shared causal loci) is more suited for identifying shared genetic architecture.

A second contribution is the multi-omics approach. Instead of relying on a single gene mapping approach, we triangulated evidence across physical proximity (MAGMA) and genetically regulated molecular traits (TWAS, spTWAS, PWAS). This is particularly valuable because it allows the data to indicate whether shared/differential signals are more consistent with expression-, splicing-, and/or protein-level mechanisms.

Notably, previous studies have rarely investigated shared gene-level signals simultaneously across expression, splicing, and protein levels; to our knowledge, there are no similar studies to the present work in respiratory disorder biology. Current colocalization algorithms mostly operate at the SNP level, which might have limited ability to map share signals at other omics layers. On the other hand, the cofdr algorithm employed in this study is flexible and can accept vectors of P-values of Z-scores, including those from other molecular-level analysis (e.g. TWAS, spTWAS, PWAS). It can also accept multiple traits (>2) as input, further enhancing the ability to detect shared signals.

Finally, we incorporated multi-trait conditioning (mtCOJO) as an additional approach to highlight genes specific to COVID-19. Conditioning COVID-19 GWAS on multiple respiratory traits provides a way to identify signals that remain associated with COVID-19 even after accounting for other related respiratory conditions.

### Revealing “differential” signals that distinguish COVID-19 from other respiratory diseases

A key novel contribution of this work is the identification of a refined set of biologically coherent “differential” signals that distinguish COVID-19 from other respiratory diseases through a systematic genome-wide scan, beyond what can be inferred from global genetic correlation or shared genome-wide significant loci. Across pairwise comparisons, we detected 214 differential loci, including 48 loci with opposite allelic directions between COVID-19 and a comparator trait. Our findings highlighted not only shared susceptibility but also genetic trade-offs (protective for one disorder but deleterious for another) across COVID-19 and other respiratory conditions. Below we highlight some of our important findings.

### Pathways distinguishing COVID-19 from other respiratory infections

A key question regarding COVID-19 pathology is: *What are the unique mechanisms that drive COVID-19 pathophysiology when compared to similar acute respiratory infections such as pneumonia and influenza?* Using a case-case GWAS approach, our differential loci (DDx) and corresponding gene-set analysis (MAGMA-DDx) isolated genomic signals that are more strongly associated with SARS-CoV-2 than other respiratory infections (Table 7 Panel A; Supplementary Table 4.7).

#### Dysregulated Type I and Type III interferon responses underlying severe COVID-19

Across comparisons of COVID vs. pneumonia and COVID vs. influenza, the most recurrent enrichments converge on interferon biology, including BIOCARTA_IFNA_PATHWAY, REACTOME_INTERFERON_ALPHA_BETA_SIGNAL ING, REACTOME_REGULATION_OF_IFNA_IFNB_SIGNALING, GOBP_INTERFERON_MED IATED_SIGNALING_PATHWAY, and GOBP/WP_TYPE_III_INTERFERON_SIGNALING.

These findings suggest the genetic architecture of severe COVID-19 is defined by a specific dysregulation of Type I and Type III interferon responses, concordant with previous reports which highlighted the importance of these pathways^66–69^. Importantly, interferons have also been investigated in clinical trials. Type III interferons, specifically peginterferon lambda-1a, showed promising results in a recent randomized controlled trial^64^. In predominantly vaccinated outpatients, a single subcutaneous dose within 7 days of symptom onset reduced the risk of hospitalization or emergency room visits by ∼51%. Our work provides further support to interferon dysregulation in COVID-19 based on human genetics evidence, suggesting that human genetic data may shed light on drug discovery or repurposing.

In summary, our DDx analysis indicates dysfunctional interferon-mediated (Type I/III) antiviral response is a potential defining genetic mechanism underlying severe COVID-19 compared to other respiratory infections. These pathway enrichments also provide functional context for the strong splicing-based differential signals at interferon-related genes (e.g., *IFNAR2* in spTWAS-DDx).

#### Alveolar epithelial, macrophage, and surfactant biology in COVID-19

A second converging theme in the COVID-versus-pneumonia/flu differential pathway results is alveolar epithelial and surfactant biology. Related pathways included, for example, GOBP_SURFACTANT_HOMEOSTASIS, GOCC_ALVEOLAR_LAMELLAR_BODY, and REACTOME_DISEASES_ASSOCIATED_WITH_SURFACTANT_METABOLISM. These enrichments highlight the role of type II alveolar epithelial cell function and the surfactant system in COVID-19, consistent with the clinical and pathological findings of diffuse alveolar injury in severe COVID-19^70, 71^.

Additionally, we identified “*Defective CSF2RB causes Pulmonary Surfactant Metabolism Dysfunction (SMDP5)*” as a pathway showing enrichment in the differential analysis, which suggests failure of surfactant homeostasis and alveolar macrophage dysfunction as a COVID-specific mechanism. *CSF2RB* is the receptor for GM-CSF, a cytokine critical for the maturation of alveolar macrophages and their ability to clear surfactants from the lungs^72^. Our findings suggest that genetic predisposition to severe COVID-19 is partially due to a distinct failure in the GM-CSF/alveolar macrophage axis, leading to the accumulation of cellular debris and surfactant dysregulation, also typical of acute respiratory distress syndrome (ARDS).

Notably, GM-CSF-based treatments (administration or inhibition) have been tested in clinical trials for respiratory infections including COVID-19. Some trials showed protective effects for COVID-19^73, 74^, though the overall results were mixed^75^. Our study provides genetic evidence that GM-CSF may indeed be a promising and specific target for COVID-19 when compared to other respiratory infections.

### Differential tissue involvement in COVID-19 versus other respiratory infections

Our tissue enrichment analysis of differential GWAS revealed that genes specifically associated with hospitalized COVID-19 (B2), but not with pneumonia or influenza, were enriched for expression in the lung and spleen (Table 7 Panel B). While lung involvement is expected, the spleen-specific enrichment is notable. Studies have shown that splenic white pulp, responsible for immune cell production, is significantly reduced in COVID-19 patients^76^. Another study has also revealed a high rate of apoptosis of immune cells in the spleen in COVID-19^77^. Our work provides genetic support for clinical observations of splenic pathology in COVID-19 patients, suggesting such pathology may be more specific to COVID-19 compared to other respiratory infections. This tissue-specific signature may reflect the ability of SARS-CoV-2 to cause systemic immune dysregulation beyond the respiratory tract.

### *FYCO1* and *HCN3* as potential COVID-19-specific risk genes

Our multi-trait conditional analysis (mtCOJO) further identified genetic variants that influence COVID-19 risk independently of shared respiratory disease biology. Our analysis revealed several genes, including *FYCO1* and *HCN3*, as tentative COVID-19-specific genes whose association with critical COVID-19 (A2) became more significant after conditioning on other respiratory disorders (Table 6 Panel B).

*FYCO1* is a protein essential for autophagosome transport along microtubules. Although the gene is located within a known COVID-19 risk locus (3p21.31), our conditional analysis reveals that this gene might be specific to COVID-19 pathology. This suggests that autophagy pathways may play a key role in mediating COVID-19 risks.

*HCN3* (hyperpolarization-activated cyclic nucleotide-gated channel 3) is the only gene that remains significant in TWAS after conditioning, with a stronger effect size after adjustment. This gene encodes a voltage-gated ion channel, largely studied in cardiac and neuronal tissues. Interestingly, HCN channels have recently been implicated in the neuronal regulation of smooth muscle tone of the airway^78^.

### Alternative splicing: an understudied mechanism underlying shared and distinct genetic architecture

Alternative splicing has been proposed as an important mechanism underlying human diseases, but it was relatively understudied in respiratory disorders, including COVID-19. An exception was a recent work by Nakanishi *et al.*^30^, who utilized Mendelian randomization to establish the causal role of splicing in genes such as *NPNT*, *ATP11A*, *OAS1*, and *DPP9* in severe COVID-19. As a secondary analysis, they also searched for the effects of the COVID-associated splicing QTLs on other diseases using the Open Targets Genetics database. However, the scope and objectives of our study differ significantly from Nakanishi *et al.* Here, we conducted a systematic genome-wide scan of shared and distinct genetic architecture of COVID-19 with a wide array of respiratory disorders, using a multi-omics framework including the study of alternative splicing.

#### Shared alternative splicing signals

Notably, our genome-wide splicing TWAS shared-signal scan (spTWAS-cofdr) identifies additional splicing-associated genes not previously reported (Table 8 Panel A). Among the shared-signal results, the most consistent COVID-asthma signals are not limited to classical antiviral loci but include a cluster of genes with strong shared splicing association in lung and/or whole blood, such as *MED24*, *INTS12*, *TCF19*, *NCOR1*, *ZSWIM7*, *BPTF*, and *CCHCR1*, implicating the role of transcriptional control, epigenetic remodeling, and genomic stability in the shared genetic architecture of COVID-19 and asthma. Specifically, *MED24* and *INTS12* serve as bridges between signaling and RNA production^79, 80^; *NCOR1* restricts macrophage-mediated inflammation^81^; and *ZSWIM7* regulates homologous recombination and DNA repair^82^.

**Table 8.**
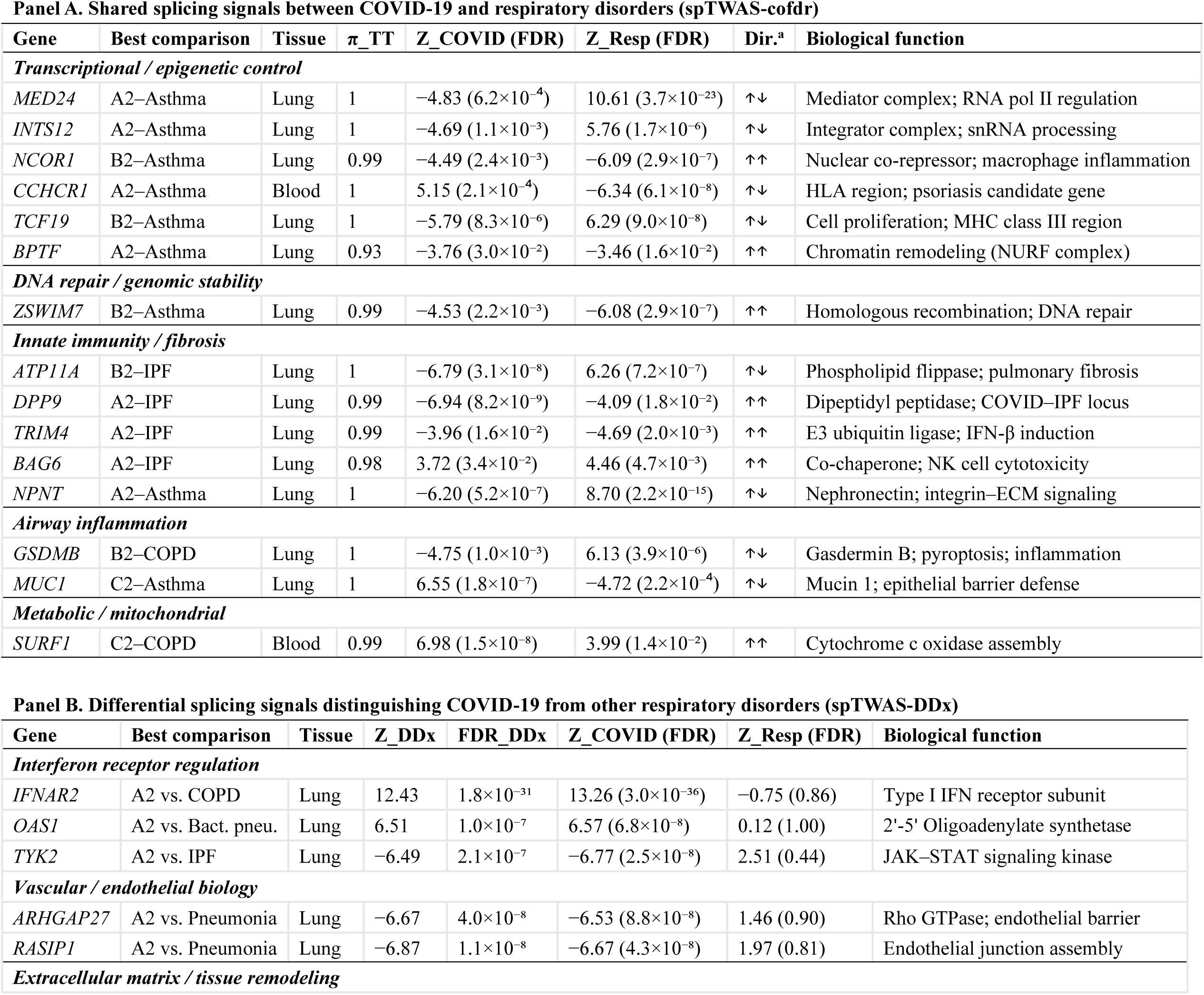
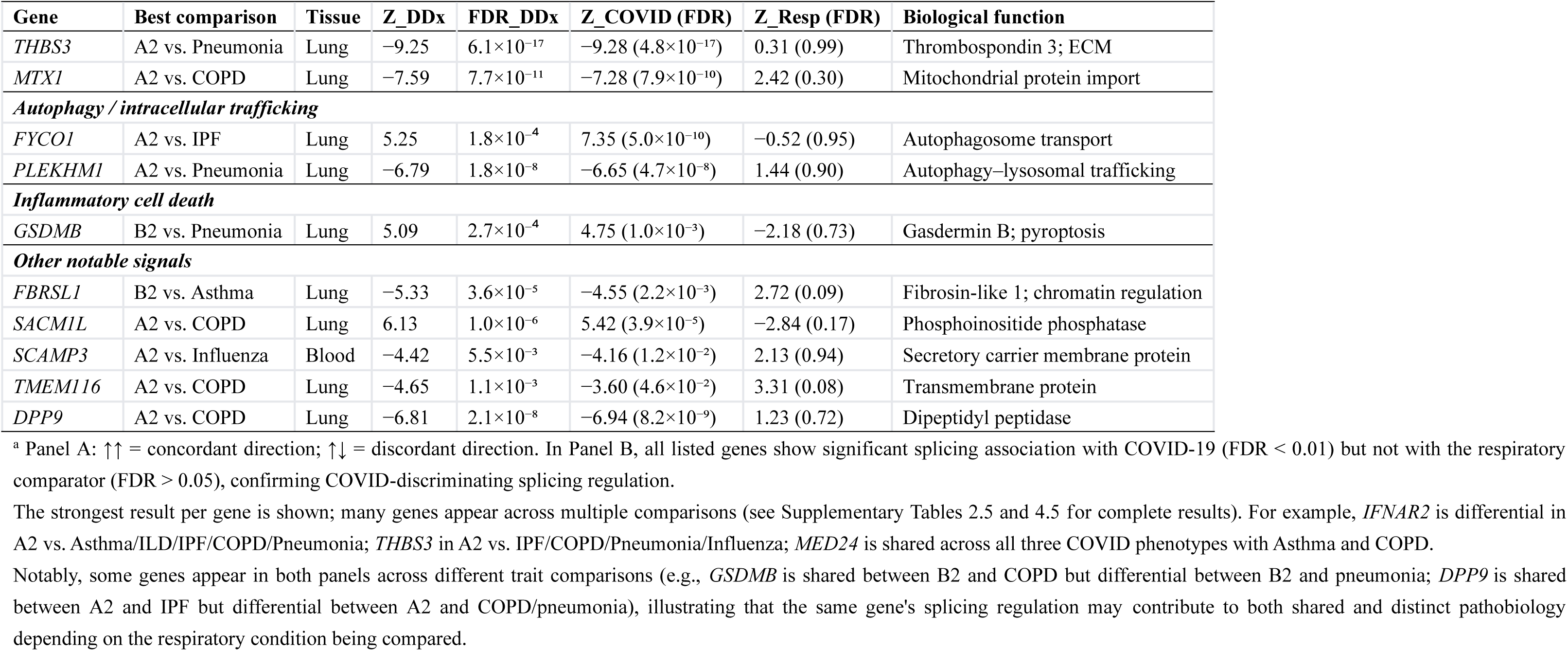
Alternative splicing signals shared with or differentiating COVID-19 and respiratory disorders.

Our shared splicing signals for COVID-IPF extend beyond the well-studied *ATP11A/DPP9* axis by highlighting additional candidates such as *TRIM4* and *BAG6*. *TRIM4* has been reported to regulate virus-induced interferon pathways^83, 84^, while *BAG6* may play a role in immune regulation by regulating NK cell activities^85^. These signals suggest that the COVID-IPF relationship may reflect shared or antagonistic regulation of innate immune responses and antigen presentation, potentially associated with hyperinflammatory responses in both diseases.

Shared splicing analysis also revealed signals connecting COVID-19 outcomes to obstructive airway disease. For example, *GSDMB* splicing was highlighted in COVID hospitalization(B2)-COPD comparisons. Notably, *GSDMB* was reported to mediate inflammatory responses in the airway following viral infection^86^.

#### COVID-discriminating splicing signals - interferon receptor regulation, vascular biology, and intracellular trafficking

Our differential spTWAS (spTWAS-DDx) results identify genes whose genetically regulated splicing signals separate COVID-19 from other respiratory disorders (Table 8 Panel B). The most prominent example is *IFNAR2*, which shows differential splicing signals across multiple comparisons (e.g., A2 vs. asthma/ILD/IPF/COPD and A2 vs. pneumonia-meta). These findings provide evidence that interferon receptor-related regulatory differences, including via alternative splicing, may be a discriminator between severe COVID-19 and other respiratory conditions.

In COVID vs. pneumonia comparisons, differential splicing involved genes such as *ARHGAP27*, *RASIP1* and *PLEKHM1*. These genes are connected to vascular/endothelial barrier function and autophagy-lysosomal biology^87, 88^. Additional differential candidates such as *MTX1* and *THBS3* (seen across multiple comparisons) further suggest roles for mitochondrial/organellar stress responses and extracellular matrix/repair programs in distinguishing COVID-19 from other respiratory conditions.

### Shared genetic signals across COVID-19, asthma, and COPD

While previous studies have focused on the genetic overlap between fibrotic lung disease and COVID-19, our systematic pairwise and multi-trait colocalization spanning eight respiratory conditions revealed a broader “pan-respiratory” genetic backbone underlying airway diseases. In particular, we identified a distinct set of genes that demonstrate pleiotropy across COVID-19, asthma, and COPD (Table 4; Supplementary Table 2.3-2.5). These genes involve several different biological pathways, such as pyroptosis and inflammatory cell death (e.g., *GSDMA*, *GSDMB*)^89^, antigen presentation and MHC signaling (*TCF19*, *MICB*)^90^, leukocyte migration and cell adhesion (*ICAM1*, *ICAM5*)^91^, and protein degradation (*PSMD3*)^92^. Our findings provide important insight into possible shared pathogenesis between COVID-19 and other chronic airway diseases.

Due to space constraints, this discussion highlights only a subset of the shared and differential splicing signals (as alternative splicing is relatively understudied), alongside selected genes/pathways with high scientific or clinical relevance. However, our study encompasses a comprehensive, genome-wide multi-omics analysis (MAGMA, TWAS, spTWAS, and PWAS) across more than 20 trait comparisons (with totally >100 sets of different analysis). The complete results are detailed in the Supplementary Tables, which we believe will serve as a valuable resource for the community to further explore the unique and shared genetic architectures, molecular mechanisms, and potential therapeutic targets underpinning COVID-19 and related respiratory phenotypes.

### Clinical implications and translational potential

Our differential analysis highlights specific genes, pathways, and pathological mechanisms underlying COVID-19 as compared to other respiratory infections, offering directions for precision medicine. First, the isolation of COVID-specific pathways, including the GM-CSF/surfactant dysfunction axis and Type III interferon signaling, suggests therapeutic approaches other than general anti-inflammatory treatments. Importantly, drugs targeting these specific mechanisms (e.g., GM-CSF and peginterferon lambda) have already entered randomized controlled trials, as discussed above. This supports the translational potential of our findings in drug discovery or repurposing. Further studies of other COVID-specific signals may be warranted.

Second, the opposing effects observed in our splicing and other omics analyses may be relevant to the side-effects of medications. For example, a drug that improves COVID-19 may theoretically lead to adverse effects on other respiratory conditions, if the drug acts on genes with opposing effects. An example is *ATP11A* in COVID-19 versus IPF, which demonstrates opposite effects in our spTWAS. Nevertheless, these hypotheses will require further testing and experimental validation.

Third, identifying shared genetic risk factors provides a foundation for the development of biomarkers for long-term sequelae. Individuals harboring risk alleles at these shared loci or showing associated changes in gene expression or protein levels, may be at heightened risk for both acute COVID-19 severity and pulmonary sequelae. As such, our findings also advocate for development of integrative polygenic risk scores (PRS) that simultaneously account for COVID-19 severity and susceptibility to respiratory comorbidities. Such scores could facilitate personalized risk stratification, forecasting not only the severity of the acute infection but also the long-term probability of post-COVID respiratory sequelae.

### Limitations and future directions

Several limitations are worth noting. Firstly, the analyses prioritize associated loci/genes and mechanistic hypotheses but do not directly establish causal genes or causal biological mechanisms. Further follow-up analysis, particularly experimental and functional validation, is required to confirm our findings. Secondly, several respiratory traits have relatively moderate case numbers, which may reduce the statistical power for both shared and differential analyses. The current study is primarily focused on the European population, and studies on more diverse ethnic groups are warranted. Finally, heterogeneity across cohorts may influence the detection of shared or differential loci and should be explored via further replication in other cohorts.

## Conclusion

In conclusion, we present a genome-wide, multi-omics and statistically rigorous framework that systematically studies shared and disease-discriminating genetic architecture between COVID-19 outcomes and multiple respiratory disorders. Our analyses highlight COVID-discriminating pathways such as interferon signaling, alveolar epithelial/surfactant biology, and GM-CSF linked mechanisms. We also prioritized COVID-specific candidate genes using conditional analyses (e.g. *FYCO1* and *HCN3*). In addition, we uncovered shared genetic signals between COVID-19 and a variety of other respiratory disorders. Together, these results shed light on the distinct and shared biological mechanisms underlying COVID-19 with other respiratory disorders, and provide an important resource of loci, genes, and pathways for follow-up functional studies, drug prioritization, and future polygenic risk prediction efforts.

## Data Availability

All data produced in the present study are available upon reasonable request to the authors

## Code availability

Custom code implementing the DDx-cofdr workflow and downstream analyses is available at: https://github.com/xxxue96/ddx_cofdr_workflow

## Acknowledgements

This work was partially supported by the KIZ-CUHK Joint Laboratory of Bioresources and Molecular Research of Common Diseases, the Hong Kong Branch of the Chinese Academy of Sciences Center for Excellence in Animal Evolution and Genetics, the Lo-Kwee Seong Biomedical Research Fund from CUHK, and a National Natural Science Foundation of China grant (HCS, grant number 81971706).

A preprint of this manuscript has been posted on medRxiv. The present submission represents the same work and has not been published elsewhere in a peer-reviewed journal.

During the preparation of this manuscript, the authors used ChatGPT (OpenAI), Claude and Grammarly to improve language and readability. All content was subsequently reviewed and edited by the author, who takes full responsibility for the accuracy and integrity of the work.

